# *In Vivo* and Phantom Repeatability of Diffusion-Weighted MRI Sequences on 1.5T MRI-Linear Accelerator (MR-Linac) and MR Simulator Devices for Head and Neck Cancers: Results from a Prospective R-IDEAL Stage 2a Evaluation of Tumor and Normal Tissue Apparent Diffusion Coefficients as Quantitative Imaging Biomarkers

**DOI:** 10.1101/2022.05.28.22275724

**Authors:** Brigid A. McDonald, Travis Salzillo, Samuel Mulder, Sara Ahmed, Alex Dresner, Kathryn Preston, Renjie He, John Christodouleas, Abdallah S. R. Mohamed, Marielle Philippens, Petra van Houdt, Daniela Thorwarth, Jihong Wang, Amita Shukla Dave, Michael Boss, Clifton D. Fuller

**Affiliations:** The University of Texas MD Anderson Cancer Center; Philips Healthcare; Elekta AB; University Medical Center Utrecht; National Cancer Institute; University of Tuebingen; Memorial Sloan Kettering Cancer Center; American College of Radiology

## Abstract

**Introduction:** Diffusion-weighted imaging (DWI) on MRI-linear accelerator (MR-linac) systems can potentially be used for monitoring treatment response and adaptive radiotherapy in head and neck cancers (HNC) but requires extensive validation. We perform technical validation to compare six total DWI sequences on an MR-linac and MR simulator (MR sim) in patients, volunteers, and phantoms.

**Methods:** Ten human papillomavirus-positive oropharyngeal cancer patients and ten volunteers underwent DWI on a 1.5T MR-linac with three DWI sequences: echo planar imaging (EPI), split acquisition of fast spin echo signals (SPLICE), and turbo spin echo (TSE). Volunteers were also imaged on a 1.5T MR sim with three sequences: EPI, BLADE, and RESOLVE. Participants underwent two scan sessions per device and two repeats of each sequence per session. Repeatability and reproducibility within-subject coefficient of variation (wCV) of mean ADC were calculated for tumors and lymph nodes (patients) and parotid glands (volunteers). Differences in measured ADC values between sequences were quantified using Bland-Altman analysis. ADC bias, repeatability/reproducibility metrics, and SNR were quantified using a phantom.

**Results:** *In vivo* repeatability/reproducibility wCV of mean ADC for parotids were 5.41%/6.72%, 3.83%/8.80%, 5.66%/10.03%, 3.44%/5.70%, 5.04%/5.66%, 4.23%/7.36% for EPI_MR-linac_, SPLICE, TSE, EPI_MR sim_, BLADE, RESOLVE. Repeatability/reproducibility wCV for EPI_MR-linac_, SPLICE, TSE were 9.64%/10.28%, 7.84%/8.96%, 7.60%/11.68% for tumors and 7.80%/9.95%, 7.23%/8.48%, 10.82%/10.44% for nodes. Bland-Altman analysis revealed significant differences between all sequence pairs except BLADE-EPI_MR-linac_ and RESOLVE-SPLICE. All sequences except TSE had phantom ADC biases within ±0.1×10^−3^ mm^2^/s for most vials. MR-linac sequences had inconsistent ADC values between different vials with the same known ADC value, indicating spatial inhomogeneities. SNR of b=0 images was 87.3, 180.5, 161.3, 171.0, 171.9, 130.2 for EPI_MR-linac_, SPLICE, TSE, EPI_MR sim_, BLADE, RESOLVE.

**Conclusion:** MR-linac DWI sequences demonstrate near-comparable performance to MR sim sequences and warrant further clinical validation for treatment response assessment in HNC.

## Introduction

Diffusion-weighted imaging (DWI) is a quantitative magnetic resonance imaging (MRI) technique that measures diffusion of water molecules in tissue, a surrogate of tissue cellularity. DWI has many applications for head and neck cancer (HNC) imaging, including lesion characterization and prediction and treatment response assessment [1–5]. Several recent studies have focused on understanding how serial DWI throughout chemotherapy and/or radiotherapy (RT) can be used to monitor response and adapt treatments based on individual response [5–12]. However, longitudinal imaging is burdensome to patients and clinicians and is generally infeasible outside of specialized research studies.

The clinical implementation of hybrid MRI/linear accelerator (MR-linac) devices has made it possible to acquire quantitative MRI sequences during every RT treatment fraction [13–17]. Current MR-linac systems enable on-line treatment plan adaptation based on changes in tumor size and shape and anatomical deformations. With further software development and validation of quantitative MRI sequences, biological image-guided adaptive RT on MR-linac systems may soon become a clinical reality [18,19].

Still, hardware modifications of current MR-linac systems to accommodate linear accelerator integration introduce additional challenges for acquiring robust quantitative MRI information. The 1.5T MR-linac employs a split gradient coil design to allow radiation beam passage, which may contribute to magnetic field gradient non-linearities [20,21]. The maximum gradient strength and slew rate of this system are lower than conventional MRIs, which necessitates longer diffusion times for the same b-value and reduces the signal-to-noise ratio (SNR) [20]. The radiolucent 2×4 channel body coil array also reduces SNR compared to other commonly used coils [22]. In light of these challenges, the MR-Linac Consortium has released guidelines for acquiring DWI on this system [20], which has informed the selection of sequence parameters in this study.

In RT, spatial accuracy of images is crucial to ensure the precise delivery of radiation. Single-shot echo planar imaging (EPI), the most commonly used readout method for DWI, is prone to severe geometric distortions and susceptibility artifacts, especially in the head and neck [23]. Turbo spin echo (TSE)-based DWI sequences have been shown to improve spatial fidelity [23,24] and are of interest for biological image-guided adaptive RT applications. However, destructive interference between spin echoes and stimulated echoes can reduce SNR in TSE-DWI. An alternative TSE-based method, “split acquisition of fast spin echo signals” (SPLICE), acquires the spin echo and stimulated echo contributions separately to preserve SNR while maintaining the spatial accuracy of TSE [25,26].

In this study, we investigate the performance of EPI, TSE, and SPLICE DWI sequences on the 1.5T MR-linac and compare them to three DWI sequences on a 1.5T diagnostic-quality MR simulation (MR sim) scanner. The MR sim sequences include EPI and two additional low-distortion sequences: “BLADE,” which uses a radial blade k-space acquisition, and “readout segmentation of long variable echo trains” (RESOLVE), a multi-shot EPI sequence. In this R-IDEAL stage 2a^1^ study [27], we perform technical validation of these six DWI sequences using data from human papillomavirus-positive (HPV+) oropharyngeal cancer patients, healthy volunteers, and a diffusion phantom.

## Methods

### Participants and Imaging

Ten patients and ten healthy volunteers were included in this study. All participants provided written informed consent; patients were consented to the MOMENTUM observational clinical trial [28] and volunteers to an internal volunteer imaging protocol, both approved by MD Anderson Cancer Center’s institutional review board. Inclusion criteria for patients included non-recurrent, histologically confirmed HPV+ oropharyngeal cancer with no prior history of cancer therapy. All imaging occurred between diagnosis and the start of treatment. Clinical demographics are in Table 1.

**Table 1:** Clinical demographics for patients and healthy volunteers. All patients had non-recurrent, histologically confirmed HPV+ oropharyngeal cancer.

| Characteristic | Patients | Healthy Volunteers |
| --- | --- | --- |
| **Sex** |  |  |
| Male | 10 (100%) | 7 (70%) |
| Female | 0 (0%) | 3 (30%) |
| **Age (years)** |  |  |
| Median (range) | 61 (58-74) | 30 (24-43) |
| **Disease sub-site** |  |  |
| Tonsil | 4 (40%) |  |
| Base of tongue | 6 (60%) |  |
| **T stage** |  |  |
| T1 | 5 (50%) |  |
| T2 | 2 (20%) |  |
| T3 | 3 (30%) |  |
| **N stage** |  |  |
| N1 | 7 (70%) |  |
| N2 | 3 (30%) |  |
| **M stage** |  |  |
| M0 | 10 (100%) |  |
| **Clinical Stage** |  |  |
| I | 6 (60%) |  |
| II | 4 (40%) |  |

Patients and volunteers were imaged on a 1.5T MR-linac (Unity; Elekta AB; Stockholm, Sweden) with a 3-D fat-suppressed T2-weighted MRI sequence and three DWI sequences: EPI_MR-linac_, SPLICE, and TSE. Volunteers were also imaged on a 1.5T MR sim (MAGNETOM Aera; Siemens Healthcare; Erlangen, Germany) with a multi-slice fat-suppressed T2-weighted MRI sequence and three DWI sequences: EPI_MR sim_, BLADE, and RESOLVE. Sequence descriptions and parameters are shown in Supplementary Tables S1 and S2. The MR-linac acquisitions used a rigid radiolucent 2×4 channel array coil [29], and the MR sim acquisitions used two 4-channel flex coils. Acquisition times (minutes) were 3.07, 7.38, 4.90, 2.93, 7.13, and 6.75 for EPI_MR-linac_, SPLICE, TSE, EPI_MR sim_, BLADE, and RESOLVE, respectively.

The diffusion b-values used were 0, 150, 500 s/mm^2^ for the MR-linac, 0, 500 s/mm^2^ for EPI on the MR sim, and 0, 800 s/mm^2^ for BLADE and RESOLVE. The choice of b-values for the MR sim was based on scan protocols used in clinical trials at our institution, which were used in this study without modification.

MR-linac b-values were chosen based on the MR-Linac Consortium’s recommendations [20]. However, because only two b-values were used for the MR sim images, ADC maps for the MR-linac images were reconstructed only with the 0 and 500 s/mm^2^ images for direct comparison to the MR sim. ADC maps were reconstructed using the built-in software on each scanner. An analysis in the Supplementary Data explores differences in ADC values and repeatability/reproducibility metrics between ADC maps reconstructed with b=0,500 mm^2^/s (b_0,500_) and b=150,500 mm^2^/s (b_150,500_).

Each study participant underwent two scan sessions per device. The first and second time points occurred at least one day apart, depending on clinical scheduling availability; mean (range) number of days between scans was 8 (1-15) for patients and 6 (1-21) for volunteers. All participants were imaged in custom RT immobilization masks to minimize motion and ensure setup reproducibility. During each session, participants were scanned twice with each DWI sequence, with a short “coffee break” out of the mask between each set to test repeatability.

### ***In Vivo*** Data Analysis

A radiologist with 5 years of experience delineated the primary tumor and pathological lymph nodes (patients) and parotid glands (volunteers). One patient did not have an MR-visible primary tumor. A total of 9 primary tumors, 30 lymph nodes, and 20 parotid glands were analyzed. Regions of interest were delineated on T2-weighted images and rigidly copied to the high-b-value image of each DWI then manually edited to account for any distortion. Segmentations were rigidly copied to corresponding ADC maps.

Repeatability/reproducibility metrics (within-subject coefficient of variation (wCV) of mean ADC) were calculated for each DWI sequence and structure type according to the Quantitative Imaging Biomarker Alliance (QIBA) consensus recommendations [30,31]. 95% confidence intervals for wCV were calculated using a chi-square statistic with n(K-1) degrees of freedom, where n is the number of sets of replicate measurements and K is the number of repeats [32]. For the repeatability (i.e. short-term) wCV calculation, there were two pairs of replicate measurements per patient/volunteer (two replicate images from the first scan session and two replicate images from the second scan session). For the reproducibility (i.e. long-term) wCV calculation, there were also two pairs of replicate measurements per patient/volunteer (the first images from the first and second scan sessions were paired, and the second images from the first and second scan sessions were paired).

Bland-Altman analysis was performed between all pairs of DWI sequences to measure differences in calculated ADC values. Values from all four imaging time points were included. Mean difference (bias) values, 95% confidence intervals for the mean differences, and 95% Bland-Altman limits of agreement were calculated in JMP (v15.0.0; SAS Institute Inc.; Cary, NC, USA) using the Method Comparison add-in. Bland-Altman analysis was also performed to assess differences in ADC values between b_0,500_ and b_150,500_ ADC maps (Supplementary Data).

### Phantom Data Acquisition and Analysis

Four sequential repeats of each DWI sequence were acquired of the QIBA diffusion phantom (model 128; CaliberMRI; Boulder, CO) for ADC bias, repeatability wCV and repeatability coefficient (RC), and SNR calculations. ADC bias, wCV, RC, and SNR were calculated for each sequence using methods described in the QIBA guidelines [30,31]. ADC bias was calculated for each vial by measuring the mean ADC value in each region of interest (ROI) and subtracting the manufacturer-provided ADC. The SNR calculation involved creating a “signal image” from the voxel-wise average of the four images and a “temporal noise image” from the voxel-wise standard deviation. SNR is equal to the ROI mean for the “signal image” divided by the ROI mean for the “temporal noise image.” A single repeat of each sequence was also acquired at a separate time point for reproducibility measurements, i.e. wCV and reproducibility coefficient (RDC). ADC bias was calculated for each vial, while the other quantities were calculated only in the central phantom vial, per the guidelines.

## Results

Representative images the six DWI sequences of a patient and volunteer are shown in Figure 1. *In vivo* mean ADC values and repeatability/reproducibility wCV values are shown in Table 2. Reproducibility wCV values were higher than repeatability wCV values for all sequences and structure types. For the MR-linac sequences with which both patients and volunteers were imaged, both repeatability and reproducibility wCV values were consistently higher for tumors and nodes than for parotid glands. In parotid glands, wCV values were slightly higher overall for the MR-linac sequences compared to the MR sim sequences.

**Table 2:** *In vivo* mean ADC values and repeatability/reproducibility wCV for each DWI sequence. Volume and mean ADC values are represented as mean ± standard deviation. wCV values are represented as wCV (upper and lower 95% confidence interval) and expressed as a percentage. Sequences are color-coded to group sequences with similar acquisition mechanisms that can be directly compared.

|  |  | EPI<sub>MR-linac</sub> | SPLICE | TSE | EPI<sub>MRsim</sub> | BLADE | RESOLVE |
| --- | --- | --- | --- | --- | --- | --- | --- |
| Sequence Description |  | Single-shot SE EPI | Single-shot TSE with split acquisition of spin echo and stimulated echo | Single-shot TSE | Single-shot SE EPI | Multi-shot radial TGSE | Multi-shot SE EPI |
| **Parotid Glands**<br>Volume (cm<sup>3</sup>):<br>32.9 ± 16.0 | Mean ADC (x10<sup>-3</sup> mm<sup>2</sup>/s) | 1.14 ± 0.17 | 1.20 ± 0.23 | 0.93 ± 0.18 | 1.49 ± 0.20 | 1.14 ± 0.15 | 1.19 ± 0.16 |
|  | Repeatability wCV (%) | 5.41 (4.42 – 6.97) | 3.83 (3.13 – 4.94) | 5.66 (4.63 – 7.29) | 3.44 (2.81 – 4.43) | 5.04 (4.12 – 6.50) | 4.23 (3.46 – 5.45) |
|  | Reproducibility wCV (%) | 6.72 (5.49 – 8.66) | 8.80 (7.19 – 11.34) | 10.03 (8.20 – 12.92) | 5.70 (4.64 – 7.40) | 5.66 (4.62 – 7.29) | 7.36 (5.99 – 9.56) |
| **Primary Tumors**<br>Volume (cm<sup>3</sup>):<br>10.7 ± 10.4 | Mean ADC (x10<sup>-3</sup> mm<sup>2</sup>/s) | 1.33 ± 0.22 | 1.80 ± 0.19 | 1.36 ± 0.19 | | | |
|  | Repeatability wCV (%) | 9.64 (7.23 – 14.45) | 7.84 (5.84 – 11.93) | 7.60 (5.70 – 11.40) |  |  |  |
|  | Reproducibility wCV (%) | 10.28 (7.65 – 15.64) | 8.96 (6.56 – 14.14) | 11.68 (8.70 – 17.78) |  |  |  |
| **Lymph Nodes**<br>Volume (cm<sup>3</sup>):<br>7.3 ± 8.0 | Mean ADC (x10<sup>-3</sup> mm<sup>2</sup>/s) | 1.38 ± 0.40 | 1.53 ± 0.36 | 1.31 ± 0.42 | | | |
|  | Repeatability wCV (%) | 7.80 (6.59 – 9.55) | 7.23 (6.06 – 8.96) | 10.82 (9.15 – 13.25) |  |  |  |
|  | Reproducibility wCV (%) | 9.95 (8.37 – 12.25) | 8.48 (7.00 – 10.78) | 10.44 (8.79 – 12.86) |  |  |  |

**Figure 1:**
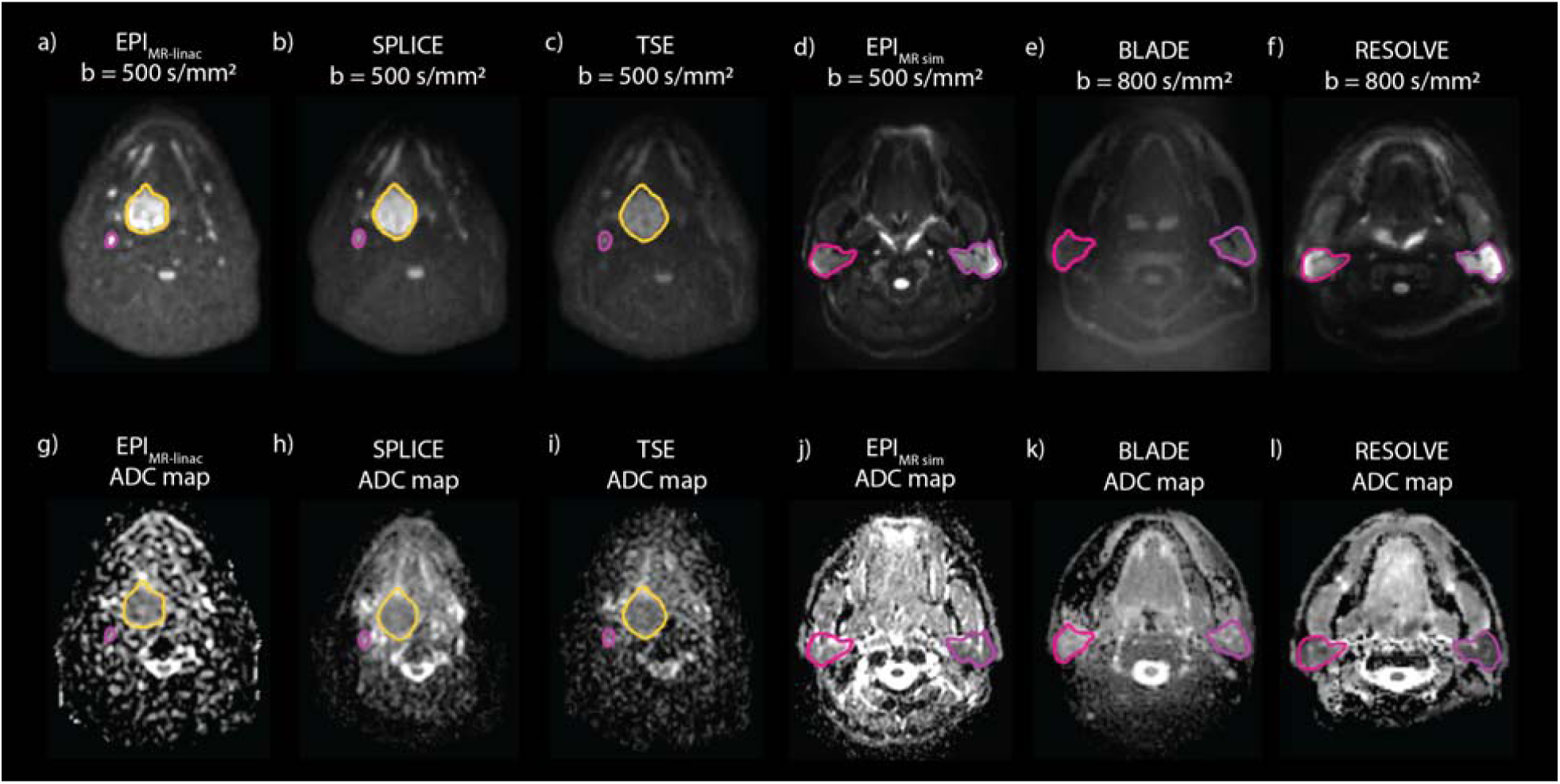
Representative high b-value images and ADC maps of a patient imaged with EPI (a, g), SPLICE (b, h), and TSE (c, i) on the MR-linac and a volunteer imaged with EPI (d, j), BLADE (e, k), and RESOLVE (f, l) on the MR sim. The primary tumor (yellow) and a lymph node (pink) are segmented on the patient images, while the parotid glands are segmented on the volunteer images.

Differences in mean ADC and wCV between b_0,500_ and b_150,500_ ADC maps are shown in Supplementary Table S3. Mean ADC values were consistently higher for b_0,500_ than for b_150,500_ ADC maps. Both repeatability and reproducibility wCV values were higher for the b_150,500_ ADC maps in nearly all cases.

Bland-Altman analysis (Figure 2, Supplementary Figure S1) revealed statistically significant biases for all pairs of DWI sequences except BLADE-EPI_MR-linac_ and RESOLVE-SPLICE. Mean differences between sequences ranged from 0.00 (BLADE-EPI_MR-linac_) to 0.56×10^−3^ mm^2^/s (EPI_MR sim_ -TSE). For the MR-linac sequences where both patient and volunteer data were compared, the SPLICE-EPI_MR-linac_ and TSE-EPI_MR-linac_ combinations showed different biases among structure types.

**Figure 2:**
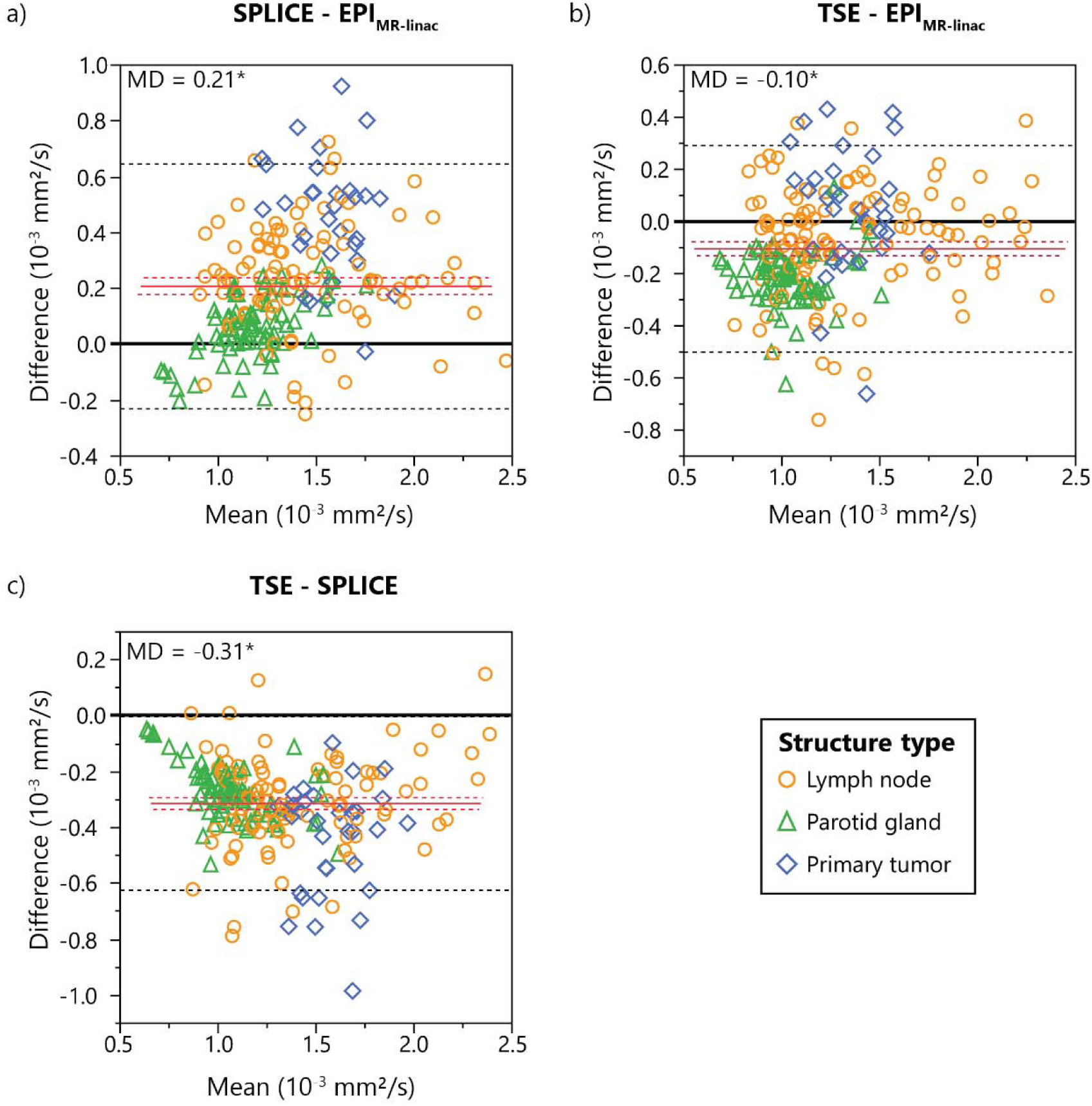
Bland-Altman plots showing differences in measured ADC values between each pair of the three MR-linac DWI sequences. Mean differences (MD) are represented by solid red lines with 95% confidence intervals of the MD represented by dotted red lines. 95% limits of agreement are represented by dotted black lines. Statistically significant biases are represented by * (p<0.05).

Bland-Altman plots showing the differences in calculated ADC values between b_0,500_ and b_150,500_ ADC maps are shown in Supplementary Figure S2. b_0,500_ overestimated b_150,500_ by 0.34, 0.48, and 0.31 ×10^−3^ mm^2^/s for EPI_MR-linac_, SPLICE, and TSE, respectively.

Phantom ADC bias results across the range of phantom ADC values are shown in Figure 3. For all MR sim sequences, the range of ADC bias values fell within ±0.1×10^−3^ mm^2^/s (except BLADE at 1.127×10^−3^ mm^2^/s). For the MR-linac sequences, EPI_MR-linac_ and SPLICE had all ADC bias values fall within ±0.1×10^−3^ mm^2^/s for the first four vials, but ADC overestimation occurred for EPI_MR-linac_ and SPLICE at the lowest two ADC values. TSE underestimated ADC by more than 0.1 mm^2^/s for nearly all vials. In general, the MR sim sequences were more precise than the MR-linac sequences. A similar graph showing the ADC bias results for each individual vial in the phantom is shown in Supplementary Figure S3. This alternative representation reveals that ADC values for the MR-linac sequences tend to be consistent for each vial across replicate images but inconsistent across different vials with the same true ADC value.

**Figure 3:**
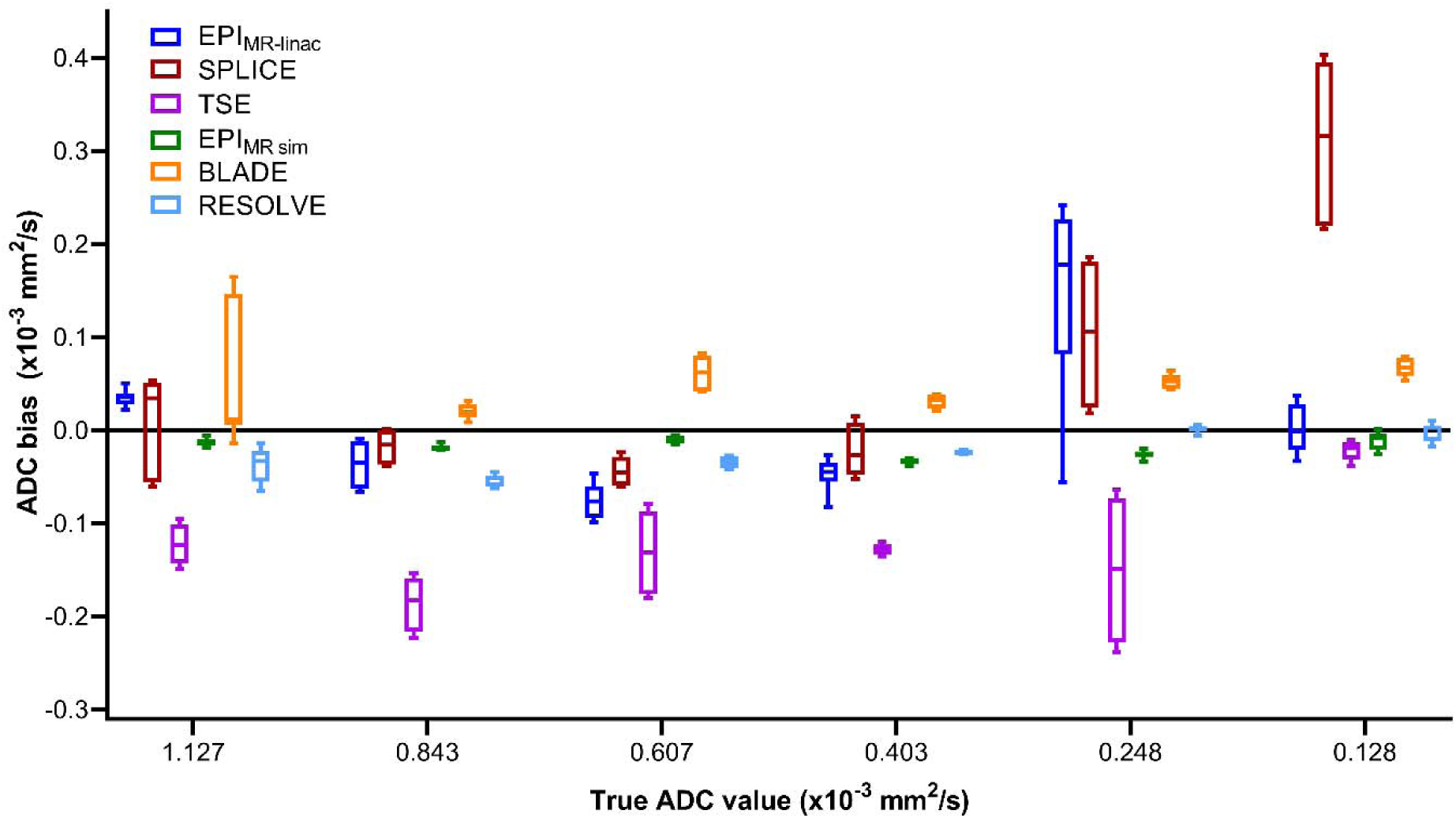
Phantom ADC bias results from replicate image acquisitions. Data points from multiple vials with the same true ADC value are combined. True ADC values are provided by the phantom manufacturer.

Phantom ADC bias (in the central vial), %CV, repeatability and reproducibility metrics, and SNR are shown in Table 3. Tolerance values from the QIBA Profile [31] are also included for reference. All sequences except SPLICE and TSE met the ±40×10^−6^ mm^2^/s criterion for ADC bias, while SPLICE and TSE had values of -57.4 and -123.0×10^−6^ mm^2^/s, respectively. For %CV, only EPI_MR sim_ and BLADE met the 2% threshold, but all MR-linac sequences were close (≤3.62%). RESOLVE had a much higher %CV (8.30%). For RC, all MR-linac sequences and EPI_MR sim_ fell under 15×10^−6^ mm^2^/s, but BLADE and RESOLVE did not (26.88 and 27.93 ×10^−6^ mm^2^/s, respectively). All sequences were within the RDC limit. All sequences exceeded the SNR threshold of 50, but EPI had the lowest SNR (87.3 for b=0).

**Table 3:** Phantom ADC bias, repeatability metrics, and SNR for each DWI sequence. ADC bias was calculated using the central phantom vial. The ADC bias and within-ROI %CV are expressed as the mean ± standard deviation of the values measured in each of the four replicate images. Tolerance values from the QIBA Profile are included in the last column for comparison.

|  | EPI (MR-linac) | SPLICE (MR-linac) | TSE (MR-linac) | EPI (MR sim) | BLADE (MR sim) | RESOLVE (MR sim) | QIBA tolerance values |
| --- | --- | --- | --- | --- | --- | --- | --- |
| ADC bias ($10^{-6} \text{ mm}^2/\text{s}$) | $26.0 \pm 3.8$ | $-57.4 \pm 3.0$ | $-123.0 \pm 1.5$ | $-15.4 \pm 2.4$ | $-0.6 \pm 9.7$ | $-23.6 \pm 10.1$ | $\leq \pm 40$ |
| %CV (within-ROI) | $2.55 \pm 0.71$ | $3.14 \pm 0.11$ | $3.62 \pm 0.30$ | $1.76 \pm 0.12$ | $1.85 \pm 0.12$ | $8.30 \pm 3.04$ | $\leq 2$ |
| RC ($\times 10^{-6} \text{ mm}^2/\text{s}$) | 10.65 | 8.15 | 4.07 | 6.67 | 26.88 | 27.93 | $\leq 15$ |
| Repeatability wCV (%) | 0.33 | 0.28 | 0.15 | 0.22 | 0.86 | 0.91 | N/A |
| RDC ($\times 10^{-6} \text{ mm}^2/\text{s}$) | 30.50 | 25.38 | 2.65 | 5.49 | 41.57 | 4.92 | $\leq 65$ |
| Reproducibility wCV (%) | 0.96 | 0.85 | 0.10 | 0.18 | 1.36 | 0.16 | N/A |
| SNR $b=0 \text{ s/mm}^2$ | 87.3 (78.6 – 96.1) | 180.5 (168.0 – 193.0) | 161.3 (146.7 – 175.9) | 171.0 (160.8 – 181.2) | 171.9 (147.5 – 196.3) | 130.2 (112.3 – 148.0) | $\geq 50 \pm 5$ |
| SNR $b=150 \text{ s/mm}^2$ | 118.9 (102.8 – 135.0) | 193.1 (183.6 – 202.6) | 266.2 (242.2 – 290.1) | N/A | N/A | N/A | N/A |
| SNR $b=500 \text{ s/mm}^2$ or $b=800 \text{ s/mm}^2$ | 106.7 (94.6 – 118.8) (b=500) | 184.0 (174.8 – 193.3) (b=500) | 261.6 (241.6 – 281.7) (b=500) | 232.2 (222.8 – 241.7) (b=500) | 104.1 (89.4 – 118.7) (b=800) | 97.3 (83.5 – 111.1) (b=800) | N/A |

## Discussion

The goals of this study were 1) to compare the performance of DWI on the 1.5T MR-linac with a 1.5T diagnostic quality MR sim and 2) to select an optimal DWI sequence for HNC on the MR-linac. To accomplish these goals, we quantified the ADC repeatability and reproducibility, ADC bias, and SNR of three DWI sequences each on a 1.5T MR-linac and a 1.5T MR sim *in vivo* and in a phantom.

ADC repeatability was previously quantified for various HNCs on diagnostic MRI systems by Paudyal et al. [35]. They measured a wCV of 2.38% for lymph nodes in a mixed cohort of 9 HNSCC patients imaged with EPI on a 3T MRI. This value is substantially lower than the wCV values measured for lymph nodes with the three MR-linac sequences in our study but is also lower than the values measured for parotid glands on the MR sim, which may be attributable to field strength, gradient, and coil hardware differences.

While the current study is the first to measure ADC repeatability/reproducibility for HNC on the MR-linac, repeatability/reproducibility of quantitative imaging biomarkers have been previously quantified on the MR-linac for other disease sites. Lawrence et al. [16] measured the ADC repeatability and reproducibility of brain tumors and healthy tissues on the MR-linac and a 1.5T diagnostic scanner. wCV values on the MR-linac were within 5% and were comparable to the diagnostic system. Kooreman et al. [15] assessed the reproducibility of intravoxel incoherent motion parameters in prostate cancer patients on the MR-linac and found the RDC of the diffusion coefficient to be 0.09×10^−3^ mm^2^/s for non-cancerous prostate and 0.44×10^−3^ mm^2^/s for tumors. For a HNC cohort on a 0.35T MR-linac, Yang et al. [17] did not explicitly calculate reproducibility metrics but found the ADC of the brainstem to be within 0.47-0.57×10^−3^ mm^2^/s across repeat imaging sessions. These studies demonstrate the robustness of quantitative imaging biomarkers on the MR-linac, suggesting their potential for longitudinal quantitative imaging and biological image-guided adaptive treatments. However, extensive validation of quantitative imaging biomarkers and standardization of scan protocols across sites is necessary for large-cohort, multi-center studies [13,19].

The QIBA Diffusion Profile [31] provides acceptability criteria for phantom metrics, which are included in Table 3 for comparison to measured values. All sequences except EPI_MR sim_ violated at least one tolerance value. However, it is important to note that these criteria are defined for system performance evaluation using an EPI sequence with specific parameters. Thus, they are not directly applicable to evaluate the sequences used in this paper but may serve as starting points for acceptability criteria based on clinical needs. In particular, the lowest ADC components of the phantom are best characterized using b=2000 s/mm^2^; use of lower maximum b-values can result in higher wCVs [36].

Our phantom data showed that in MR-linac sequences only, ADC values varied substantially across different vials with the same diffusivity but not across replicate images within the same vial, suggesting that spatial inhomogeneities are more significant for the MR-linac than the MR sim. Kooreman et al. [20] found similar results with a homogeneous diffusion phantom imaged on an MR-linac and MR sim. They attributed the differences to the split gradient coil design on the MR-linac, which is necessary to accommodate the radiation treatment beam but induces eddy currents closer to isocenter, causing magnetic field inhomogeneities.

Parotid gland wCV values were higher overall for the MR-linac compared to the MR sim, but differences were not severe and values were within clinically acceptable ranges. However, the spatial dependence of phantom ADC values was more substantial on the MR-linac, which may have implications for longitudinal *in vivo* studies as patient anatomy changes and setup uncertainty increases throughout treatment. Still, these results demonstrate that *in vivo* DWI of HNC is possible on the 1.5T MR-linac with acceptable repeatability/reproducibility and lay the foundation for future clinical studies.

Comparing the MR-linac sequences, EPI_MR-linac_ and SPLICE had more accurate phantom ADC values than TSE, with the exception of SPLICE at low ADC values. For repeatability, SPLICE had the lowest overall wCV values and highest overall SNR. Based on these data and our clinical preference for a low-distortion DWI sequence, SPLICE is the optimal sequence for HNC imaging on the MR-linac. However, a disadvantage of SPLICE is the long acquisition time (7 minutes for 3 b-values).

One surprising result is that the SNR of EPI_MR-linac_ was much lower than for SPLICE and TSE. A likely explanation is that short inversion time inversion recovery (STIR) was used for fat suppression for EPI_MR-linac_ and spectral presaturation with inversion recovery (SPIR) for SPLICE and TSE. STIR uses inversion recovery to nullify fat signal but reduces signal from all tissues, resulting in reduced SNR. SPIR uses a spectrally selective inversion pulse to improve SNR from non-fat tissues [37]. We used the consensus EPI protocol that had been distributed among MR-Linac Consortium [38] sites without modification for comparison across sites. Because no consensus SPLICE or TSE protocols existed, these sequences were optimized in-house, and we chose SPIR over STIR to maximize SNR. If SPIR is used for EPI, SNR and reproducibility would likely improve. However, poor fat suppression in EPI causes large chemical shift artifacts, while the non-suppressed fat in TSE and SPLICE sequences only shows a small chemical shift.

One limitation of this study was that clinical scheduling constraints prevented patients from undergoing two scans each on both the MR-linac and MR sim between the time of simulation and the start of treatment. While patients would provide the most ideal comparison between the devices, healthy volunteers were included to assess differences between the systems in parotid glands. However, our data reveal that the repeatability and ADC bias behavior of parotid glands differs from that of tumors and nodes, so future investigations using patients on both the MR-linac and MR sim should confirm the findings in this study. Furthermore, the most ideal comparison between the MR-linac and a diagnostic-quality scanner would be to use a 1.5T Philips MRI with the same DWI sequences, but we were limited by the MR sim and sequences available at our institution. Finally, we did not assess geometric distortion in this study, which is a major consideration for RT applications.

## Conclusion

We have assessed the repeatability/reproducibility, ADC bias, and SNR of DWI sequences on a 1.5T MR-linac and MR sim for HNC both *in vivo* and in phantoms and demonstrated near-comparable performance between the MR-linac and MR sim. These results show that the MR-linac DWI sequences are robust and worthy of further evaluation as a quantitative method of assessing treatment response in HNC.

## Data Availability

The images and segmentations used for this project will be anonymized and made available in a public data repository by the time of publication.

## Footnotes

1 R-IDEAL is an evaluation framework for radiation oncology technological advancements. Stage 2a is the “development” phase, including “technical improvements, feasibility, and safety.”

## References

[1] Payabvash S. Quantitative diffusion magnetic resonance imaging in head and neck tumors. Quant Imaging Med Surg 2018;8:1052–65. https://doi.org/10.21037/qims.2018.10.14.

[2] Salzillo TC, Taku N, Wahid KA, McDonald BA, Wang J, van Dijk L V., et al. Advances in Imaging for HPV-Related Oropharyngeal Cancer: Applications to Radiation Oncology. Semin Radiat Oncol 2021;31:371–88. https://doi.org/10.1016/j.semradonc.2021.05.001.

[3] Driessen JP, Van Kempen PMW, Van Der Heijden GJ, Philippens MEP, Pameijer FA, Stegeman I, et al. Diffusion-weighted imaging in head and neck squamous cell carcinomas: A systematic review. Head Neck 2015. https://doi.org/10.1002/hed.23575.

[4] Connolly M, Srinivasan A. Diffusion-Weighted Imaging in Head and Neck Cancer: Technique, Limitations, and Applications. Magn Reson Imaging Clin N Am 2018;26:121–33. https://doi.org/10.1016/j.mric.2017.08.011.

[5] Chang Z, Wang C. Treatment assessment of radiotherapy using MR functional quantitative imaging. World J Radiol 2015;7:1–6. https://doi.org/10.4329/wjr.v7.i1.1.

[6] Vandecaveye V, Dirix P, De Keyzer F, Op De Beeck K, Vander Poorten V, Hauben E, et al. Diffusion-weighted magnetic resonance imaging early after chemoradiotherapy to monitor treatment response in head-and-neck squamous cell carcinoma. Int J Radiat Oncol Biol Phys 2012;82:1098– 107. https://doi.org/10.1016/j.ijrobp.2011.02.044.

[7] Vandecaveye V, Dirix P, De Keyzer F, Op De Beeck K, Vander V, Roebben PI, et al. Predictive value of diffusion-weighted magnetic resonance imaging during chemoradiotherapy for head and neck squamous cell carcinoma. Eur Radiol 2010;20:1703–14. https://doi.org/10.1007/s00330-010-1734-6.

[8] Hong J, Yao Y, Zhang Y, Tang T, Zhang H, Bao D, et al. Value of magnetic resonance diffusion-weighted imaging for the prediction of radiosensitivity in nasopharyngeal carcinoma. Otolaryngol -Head Neck Surg 2013;149:707–13. https://doi.org/10.1177/0194599813496537.

[9] van der Heide UA, Houweling AC, Groenendaal G, Beets-Tan RGH, Lambin P. Functional MRI for radiotherapy dose painting. Magn Reson Imaging 2012;30:1216–23. https://doi.org/10.1016/j.mri.2012.04.010.

[10] Houweling AC, Wolf AL, Vogel W V., Hamming-Vrieze O, Van Vliet-Vroegindeweij C, Van De Kamer JB, et al. FDG-PET and diffusion-weighted MRI in head-and-neck cancer patients: Implications for dose painting. Radiother Oncol 2013;106:250–4. https://doi.org/10.1016/j.radonc.2013.01.003.

[11] Ligtenberg H, Schakel T, Dankbaar JW, Ruiter LN, Peltenburg B, Willems SM, et al. Target Volume Delineation Using Diffusion-weighted Imaging for MR-guided Radiotherapy: A Case Series of Laryngeal Cancer Validated by Pathology. Cureus 2018;D. https://doi.org/10.7759/cureus.2465.

[12] Schakel T, Peltenburg B, Dankbaar JW, Cardenas CE, Aristophanous M, Terhaard CHJ, et al. Evaluation of diffusion weighted imaging for tumor delineation in head-and-neck radiotherapy by comparison with automatically segmented 18F-fluorodeoxyglucose positron emission tomography. Phys Imaging Radiat Oncol 2018;5:13–8. https://doi.org/10.1016/j.phro.2017.12.004.

[13] Thorwarth D, Ege M, Nachbar M, Monnich D, Gani C, Zips D, et al. Quantitative magnetic resonance imaging on hybrid magnetic resonance linear accelerators!]: Perspective on technical and clinical validation. Phys Imaging Radiat Oncol 2020;16:69–73. https://doi.org/10.1016/j.phro.2020.09.007.

[14] Kooreman ES, van Houdt PJ, Nowee ME, van Pelt VWJ, Tijssen RHN, Paulson ES, et al. Feasibility and accuracy of quantitative imaging on a 1.5 T MR-linear accelerator. Radiother Oncol 2019;133:156–62. https://doi.org/10.1016/j.radonc.2019.01.011.

[15] Kooreman ES, van Houdt PJ, Keesman R, van Pelt VWJ, Nowee ME, Pos F, et al. Daily Intravoxel Incoherent Motion (IVIM) In Prostate Cancer Patients During MR-Guided Radiotherapy—A Multicenter Study. Front Oncol 2021;11:1–9. https://doi.org/10.3389/fonc.2021.705964.

[16] Lawrence LSP, Chan RW, Chen H, Keller B, Stewart J, Ruschin M, et al. Accuracy and precision of apparent diffusion coefficient measurements on a 1.5 T MR-Linac in central nervous system tumour patients. Radiother Oncol 2021;164:155–62. https://doi.org/10.1016/j.radonc.2021.09.020.

[17] Yang Y, Cao M, Sheng K, Gao Y, Chen A, Kamrava M, et al. Longitudinal diffusion MRI for treatment response assessment: Preliminary experience using an MRI-guided tri-cobalt 60 radiotherapy system. Med Phys 2016;43:1369–73. https://doi.org/10.1118/1.4942381.

[18] van Houdt PJ, Yang Y, van der Heide UA. Quantitative Magnetic Resonance Imaging for Biological Image-Guided Adaptive Radiotherapy. Front Oncol 2021;10:1–9. https://doi.org/10.3389/fonc.2020.615643.

[19] van Houdt PJ, Saeed H, Thorwarth D, Fuller CD, Hall WA, McDonald BA, et al. Integration of quantitative imaging biomarkers in clinical trials for MR-guided radiotherapy: Conceptual guidance for multicentre studies from the MR-Linac Consortium Imaging Biomarker Working Group. Eur J Cancer 2021;153:64–71. https://doi.org/10.1016/j.ejca.2021.04.041.

[20] Kooreman ES, van Houdt PJ, Keesman R, Pos FJ, van Pelt VWJ, Nowee ME, et al. ADC measurements on the Unity MR-linac – A recommendation on behalf of the Elekta Unity MR-linac consortium. Radiother Oncol 2020;153:106–13. https://doi.org/10.1016/j.radonc.2020.09.046.

[21] Tijssen RHN, Philippens MEP, Paulson ES, Glitzner M, Chugh B, Wetscherek A, et al. MRI commissioning of 1.5T MR-linac systems – a multi-institutional study. Radiother Oncol 2019;132:114–20. https://doi.org/10.1016/j.radonc.2018.12.011.

[22] Zijlema SE, Tijssen RHN, Malkov VN, Van Dijk L, Hackett SL, Kok JGM, et al. Design and feasibility of a flexible, on-body, high impedance coil receive array for a 1.5 T MR-linac. Phys Med Biol 2019;64. https://doi.org/10.1088/1361-6560/ab37a8.

[23] Verhappen MH, Pouwels PJW, Ljumanovic R, Van Der Putten L, Knol DL, De Bree R, et al. Diffusion-weighted MR imaging in head and neck cancer: Comparison between half-Fourier acquired single-shot turbo spin-echo and EPI techniques. Am J Neuroradiol 2012;33:1239–46. https://doi.org/10.3174/ajnr.A2949.

[24] Gao Y, Han F, Zhou Z, Cao M, Kaprealian T, Kamrava M, et al. Distortion-free diffusion MRI using an MRI-guided Tri-Cobalt 60 radiotherapy system: Sequence verification and preliminary clinical experience. Med Phys 2017;44:5357–66. https://doi.org/10.1002/mp.12465.

[25] Schakel T, Hoogduin JM, Terhaard CHJ, Philippens MEP. Technical Note: Diffusion-weighted MRI with minimal distortion in head-and-neck radiotherapy using a turbo spin echo acquisition method: Diffusion-weighted. Med Phys 2017;44:4188–93. https://doi.org/10.1002/mp.12363.

[26] Schick F. SPLICE: Sub-second diffusion-sensitive MR imaging using a modified fast spin-echo acquisition mode. Magn Reson Med 1997;38:638–44. https://doi.org/10.1002/mrm.1910380418.

[27] Verkooijen HM, Kerkmeijer LGW, Fuller CD, Huddart R, Faivre-Finn C, Verheij M, et al. R-IDEAL: A framework for systematic clinical evaluation of technical innovations in radiation oncology. Front Oncol 2017;7:1–7. https://doi.org/10.3389/fonc.2017.00059.

[28] van Otterloo SR de M, Christodouleas JP, Blezer ELA, Akhiat H, Brown K, Choudhury A, et al. The MOMENTUM Study: An International Registry for the Evidence-Based Introduction of MR-Guided Adaptive Therapy. Front Oncol 2020;10:1328. https://doi.org/10.3389/fonc.2020.01328.

[29] Hoogcarspel SJ, Van Der Velden JM, Lagendijk JJW, Van Vulpen M, Raaymakers BW. The feasibility of utilizing pseudo CT-data for online MRI based treatment plan adaptation for a stereotactic radiotherapy treatment of spinal bone metastases. Phys Med Biol 2014;59:7383–91. https://doi.org/10.1088/0031-9155/59/23/7383.

[30] Shukla-Dave A, Obuchowski NA, Chenevert TL, Jambawalikar S, Schwartz LH, Malyarenko D, et al. Quantitative imaging biomarkers alliance (QIBA) recommendations for improved precision of DWI and DCE-MRI derived biomarkers in multicenter oncology trials. J Magn Reson Imaging 2019;49:e101–21. https://doi.org/10.1002/jmri.26518.

[31] RSNA QIBA Diffusion-Weighted Imaging Task Force. QIBA Profile: Diffusion-Weighted Magnetic Resonance Imaging (DWI). 2019.

[32] Barnhart HX, Barboriak DP. Applications of the repeatability of quantitative imaging biomarkers: A review of statistical analysis of repeat data sets. Transl Oncol 2009;2:231–5. https://doi.org/10.1593/tlo.09268.

[33] Hoang JK, Choudhury KR, Chang J, Craciunescu OI, Yoo DS, Brizel DM. Diffusion-weighted imaging for head and neck squamous cell carcinoma: Quantifying repeatability to understand early treatment-induced change. Am J Roentgenol 2014;203:1104–8. https://doi.org/10.2214/AJR.14.12838.

[34] Rodrigues A, Loman K, Nawrocki J, Hoang JK, Chang Z, Mowery YM, et al. Establishing ADC-Based Histogram and Texture Features for Early Treatment-Induced Changes in Head and Neck Squamous Cell Carcinoma. Front Oncol 2021;11:708398. https://doi.org/10.3389/fonc.2021.708398.

[35] Paudyal R, Konar AS, Obuchowski NA, Hatzoglou V, Chenevert TL, Malyarenko DI, et al. Repeatability of Quantitative Diffusion-Weighted Imaging Metrics in Phantoms, Head-and-Neck and Thyroid Cancers: Preliminary Findings. Tomography 2019;5:15–25. https://doi.org/10.18383/j.tom.2018.00044.

[36] Boss MA, Chenevert TL, Waterton JC, Morris DM, Ragheb H, Jackson A, et al. Temperature-controlled Isotropic Diffusion Phantom with Wide Range of Apparent Diffusion Coefficients for Multicenter Assessment of Scanner Repeatability and Reproducibility. Proc. Int. Soc. Magn. Reson. Med. 22, 2014, p. 4505.

[37] Del Grande F, Santini F, Herzka DA, Aro MR, Dean CW, Gold GE, et al. Fat-suppression techniques for 3-T MR imaging of the musculoskeletal system. Radiographics 2014;34:217–33. https://doi.org/10.1148/rg.341135130.

[38] Kerkmeijer LGW, Fuller CD, Verkooijen HM, Verheij M, Choudhury A, Harrington KJ, et al. The MRI-Linear Accelerator Consortium: Evidence-Based Clinical Introduction of an Innovation in Radiation Oncology Connecting Researchers, Methodology, Data Collection, Quality Assurance, and Technical Development. Front Oncol 2016;6:215. https://doi.org/10.3389/fonc.2016.00215.

