## Supplementary Data for "*In Vivo* and Phantom Repeatability of Diffusion-Weighted MRI Sequences on 1.5T MRI-Linear Accelerator (MR-Linac) and MR Simulator Devices for Head and Neck Cancers: Results from a Prospective R-IDEAL Stage 2a Evaluation of Tumor and Normal Tissue Apparent Diffusion Coefficients as Quantitative Imag"

**DWI Sequences**

Table S1: Descriptions of each DWI sequence used in this study and brief discussion of pros and cons of each sequence.

| **Sequence** | **Image Acquisition Mechanism** | **Pros** | **Cons** |
| --- | --- | --- | --- |
| **EPI (MR-linac)** | Single-shot EPI: All of k-space is acquired in a single TR by rapidly switching the frequency-encoding gradient polarity | - Fast acquisition 🡪 minimal motion artifacts - High SNR compared to TSE-based sequences (when other factors such as number of averages and fat suppression are held constant) | - Severe geometric distortions and susceptibility artifacts due to accumulated phase errors and eddy currents - Low gradient amplitude (15 mT/m) and slew rate (65 T/m/s) on MR-linac lead to longer imaging times 🡪 increases risk of T2 blurring and potential for motion artifacts compared to EPI on MR sim |
| **SPLICE (MR-linac)** | Single-shot TSE with split echo acquisition: All of k-space is acquired in a single TR by applying a series of refocusing RF pulses and capturing the signal (magnitude only) from each spin echo and stimulated echo separately | - Reduced geometric distortion and susceptibility artifacts compared to EPI - Increased SNR compared to TSE because spin echoes and stimulated echoes are acquired separately | - Reduced SNR compared to EPI-based sequences (when other factors such as number of averages and fat suppression are held constant) |
| **TSE (MR-linac)** | Single-shot TSE: All of k-space is acquired in a single TR by applying a series of refocusing RF pulses and capturing the signal (magnitude and phase; CPMG components only) from each spin echo | - Reduced geometric distortion and susceptibility artifacts compared to EPI | - Reduced SNR compared to SPLICE due to destructive interference of spin echoes and stimulated echoes - Reduced SNR compared to EPI-based sequences (when other factors such as number of averages and fat suppression are held constant) |
| **EPI (MR sim)** | Single-shot EPI: All of k-space is acquired in a single TR by rapidly switching the frequency-encoding gradient polarity | - Fast acquisition 🡪 minimal motion artifacts - Increased SNR compared to TSE-based sequences (when other factors such as number of averages and fat suppression are held constant) - High gradient amplitude (45 mT/m) and slew rate (200 T/m/s) on MR sim lead to shorter imaging times compared to MR-linac 🡪 reduces risk of T2 blurring and potential for motion artifacts compared to MR-linac | - Severe geometric distortions and susceptibility artifacts due to accumulated phase errors and eddy currents |
| **BLADE (MR sim)** | Radial TGSE: k-space is acquired through a series of parallel lines rotating around the center of k-space, combined with a hybrid TSE/gradient echo acquisition | - Less prone to motion artifacts compared to other multi-shot methods because the radial acquisition diffuses motion throughout the whole image - Reduced specific absorption rate compared to TSE and EPI | - Longer acquisition times compared to single-shot EPI/TSE |
| **RESOLVE (MR sim)** | Multi-shot EPI: segments of k-space are acquired during a small number of TRs (5 in this sequence) using EPI (rapid gradient switching) | - Less prone to phase error accumulation and eddy currents compared to single-shot EPI 🡪 improved geometric accuracy and less risk of susceptibility artifacts compared to single-shot EPI (but not compared to TSE-based sequences) | - Longer acquisition times compared to single-shot EPI/TSE and potential for motion between shots 🡪 increased risk of motion artifacts (although this is mitigated by the use of a 2-D navigator) |

Abbreviations:

EPI = echo planar imaging

SPLICE = split acquisition of fast spin echo signals

TSE = turbo spin echo

RESOLVE = readout segmentation of long variable echo trains

TR = repetition time

RF = radiofrequency

CPMG = Carr-Purcell-Meiboom-Gill

TGSE = turbo gradient spin echo

SNR = signal-to-noise ratio

**MRI Sequence Parameters**

Table S2: Sequence parameters for the T2-weighted and DWI sequences on the MR-linac and MR sim.

|  | **T2 (MR-linac)** | **EPI (MR-linac)** | **SPLICE (MR-linac)** | **TSE (MR-linac)** | **T2 (MR sim)** | **EPI (MR sim)** | **BLADE (MR sim)** | **RESOLVE (MR sim)** |
| --- | --- | --- | --- | --- | --- | --- | --- | --- |
| Scan type | Fat-suppressed T2-weighted MRI | Single-shot SE EPI | Single-shot TSE with split acquisition of spin echo and stimulated echo | Single-shot TSE | Fat-suppressed T2-weighted MRI | Single-shot SE EPI | Multi-shot radial TGSE | Multi-shot SE EPI |
| TR (ms) | 1400 | 3613 | 3823 | 2530 | 5280 | 6000 | 5400 | 9380 |
| TE (ms) | 182 | 75 | 82 | 72 | 80 | 80 | 55 | 63/104* |
| Refocusing flip angle (degrees) | 55 | N/A | 80 | 100 | 180 | N/A | 120 | 180 |
| b-values (number of averages) | N/A | 0 (2), 150 (4), 500 (12) | 0 (4),150 (6), 500 (12) | 0 (4),150 (6), 500 (12) | N/A | 0 (2), 500 (8) | 0 (2), 800 (8) | 0 (2), 800 (8) |
| Half scan/partial Fourier factor | N/A | 0.88 | 1 | 0.8 | N/A | 0.75 | 1.5 (BLADE 150%) | 0.875 |
| Fat suppression | SPAIR | STIR | SPIR | SPIR | Spectral FS | Spectral FS | Spectral FS | Spectral FS |
| Bandwidth** Philips: WFS (pix)/BW (Hz/pix) Siemens: BW (Hz) | 0.407/533.4 | 6.082/35.7 | 0.722/300.7 | 0.320/678.2 | 300 | 1132 | 1220 | 888 |
| EPI/TSE factor | 72 (TSE) | 39 (EPI) | 42 (TSE) | 34 (TSE) | 15 (TSE) | 130 (EPI) | 15 (TSE) | 96 (EPI - 5 segments) |
| Acceleration type (factor) | SENSE (2) | SENSE (2.2) | SENSE (2) | SENSE (2) | None | GRAPPA (2) | None | GRAPPA (2) |
| Phase encode direction | AP | AP | AP | AP | AP | AP | N/A (radial) | AP |
| Field of view (RL x AP x FH) (mm) | 520 x 270 x 200 | 300 x 300 x 100 | 288 x 252 x 100 | 288 x 252 x 100 | 256 x 256 x 240 | 256 x 256 x 96 | 256 x 256 x 96 | 256 x 256 x 192 |
| Acquisition pixel size (RL x AP x FH) (mm) | 1.4 x 1.5 x 2 | 3.5 x 3.5 x 4 | 3 x 3 x 5 | 3 x 3 x 5 | 1 x 1 x 2 | 1.97 x 1.97 x 4 | 2 x 2 x 4 | 2 x 2 x 4 |
| Reconstruction pixel size (RL x AP x FH) (mm) | 0.81 x 0.81 x 1 | 1.6 x 1.6 x 4 | 1.5 x 1.5 x 5 | 1.5 x 1.5 x 5 | 0.81 x 0.81 x 2 | 1 x 1 x 4 | 1 x 1 x 4 | 2 x 2 x 4 |
| Total scan duration (min:sec) | 5:09 | 3:04 | 7:23 | 4:54 | 6:22 | 2:56 | 7:08 | 6:45 |

*Philips and Siemens report bandwidth differently. We report the vendor-specified values for easy reproducibility of sequences.

**Double refocused, so two TE's

Abbreviations:

EPI = echo planar imaging

SPLICE = split acquisition of fast spin echo signals

TSE = turbo spin echo

RESOLVE = readout segmentation of long variable echo trains

SE = spin echo

TGSE = turbo gradient spin echo

TR = repetition time

TE = echo time

STIR = short inversion time inversion recovery

SPIR = spectral presaturation with inversion recovery

FS = fat saturation

WFS = water-fat shift

BW = bandwidth

Pix = pixel

SENSE = sensitivity encoding

GRAPPA = generalized autocalibrating partial parallel acquisition

AP = anterior-posterior

RL = right-left

FH = foot-head

Min = minutes

Sec = seconds

**Repeatability and Reproducibility Metrics for b=0,500 vs. b=150,500 MR-Linac Sequences**

Repeatability and reproducibility metrics were calculated for ADC maps reconstructed with b = 150 and 500 s/mm^2^ (b_150,500_) and compared with those reconstructed with b = 0 and 500 s/mm^2^ (b_0,500_). To test whether the b-values used in the ADC reconstruction impacted the repeatability/reproducibility, a Wilcoxon signed rank test was performed using the wCV^2^ values for each structure calculated from the b_0,500_ and b_150,500_ ADC maps. Separate Wilcoxon signed rank tests were performed for parotid glands, primary tumors, and lymph nodes and for short-term and long-term wCV^2^. Analyses were performed in JMP.

Table S3: *In vivo* repeatability and reproducibility for MR-linac DWI sequences with ADC maps reconstructed using b = 0 and 500 s/mm^2^ (b_0,500_) vs. ADC maps reconstructed with b = 150 and 500 s/mm^2^ (b_150,500_). Mean ADC values are reported as mean ± standard deviation. wCV values are reported as the wCV with 95% confidence intervals in parentheses. Pairs of b=0,500 and b=150,500 ADC maps with a statistically significant difference in the Wilcoxon signed rank test are denoted with * under both the b_0,500_ and b_150,500_ columns.

|  |  | EPI_MR-linac_ b_0,500_ | EPI_MR-linac_ b_150,500_ | SPLICE b_0,500_ | SPLICE b_150,500_ | TSE b_0,500_ | TSE b_150,500_ |
| --- | --- | --- | --- | --- | --- | --- | --- |
| Parotid glands | Mean ADC (x10^-3^ mm^2^/s) | 1.14 ± 0.17 | 0.72 ± 0.14 | 1.20 ± 0.23 | 0.71 ± 0.18 | 0.93 ± 0.18 | 0.62 ± 0.14 |
|  | Repeatability wCV (%) | 5.41 (4.42 – 6.97) | 4.64 (3.80 – 5.99) | 3.83 (3.13 – 4.94) | 4.96 (4.05 – 6.39) | 5.66 (4.63 – 7.29)* | 6.53 (5.34 – 8.41)* |
|  | Reproducibility wCV (%) | 6.72 (5.49 – 8.66) | 6.94 (5.67 – 8.95) | 8.80 (7.19 – 11.34) | 10.02 (8.19 – 12.91) | 10.03 (8.20 – 12.92) | 9.11 (7.44 – 11.74) |
| Primary tumors | Mean ADC (x10^-3^ mm^2^/s) | 1.33 ± 0.22 | 0.96 ± 0.27 | 1.80 ± 0.19 | 1.16 ± 0.18 | 1.36 ± 0.19 | 0.96 ± 0.19 |
|  | Repeatability wCV (%) | 9.64 (7.23 – 14.45) | 22.40 (16.81 – 33.58) | 7.84 (5.84 – 11.93)* | 15.28 (11.38 – 23.25)* | 7.60 (5.70 – 11.40) | 14.72 (11.04 – 22.06) |
|  | Reproducibility wCV (%) | 10.28 (7.65 –15.64)* | 18.97 (14.13 – 28.87)* | 8.96 (6.56 – 14.14)* | 15.24 (11.16 – 24.04)* | 11.68 (8.70 – 17.78) | 14.51 (10.81 – 22.09) |
| Lymph nodes | Mean ADC (x10^-3^ mm^2^/s) | 1.38 ± 0.40 | 1.11 ± 0.41 | 1.53 ± 0.36 | 1.12 ± 0.37 | 1.31 ± 0.42 | 1.02 ± 0.41 |
|  | Repeatability wCV (%) | 7.80 (6.59 – 9.55) | 13.15 (11.11 – 16.09) | 7.23 (6.06 – 8.96)* | 9.54 (8.00 – 11.84)* | 10.82 (9.15 – 13.25)* | 13.12 (11.09 – 16.06)* |
|  | Reproducibility wCV (%) | 9.95 (8.37 – 12.25)* | 14.27 (12.01 – 17.57)* | 8.48 (7.00 – 10.78)* | 9.77 (8.06 – 12.42)* | 10.44 (8.79 – 12.86)* | 13.83 (11.64 – 17.03)* |

**Bland-Altman Analysis for all MR-Linac and MR Sim DWI Sequences**

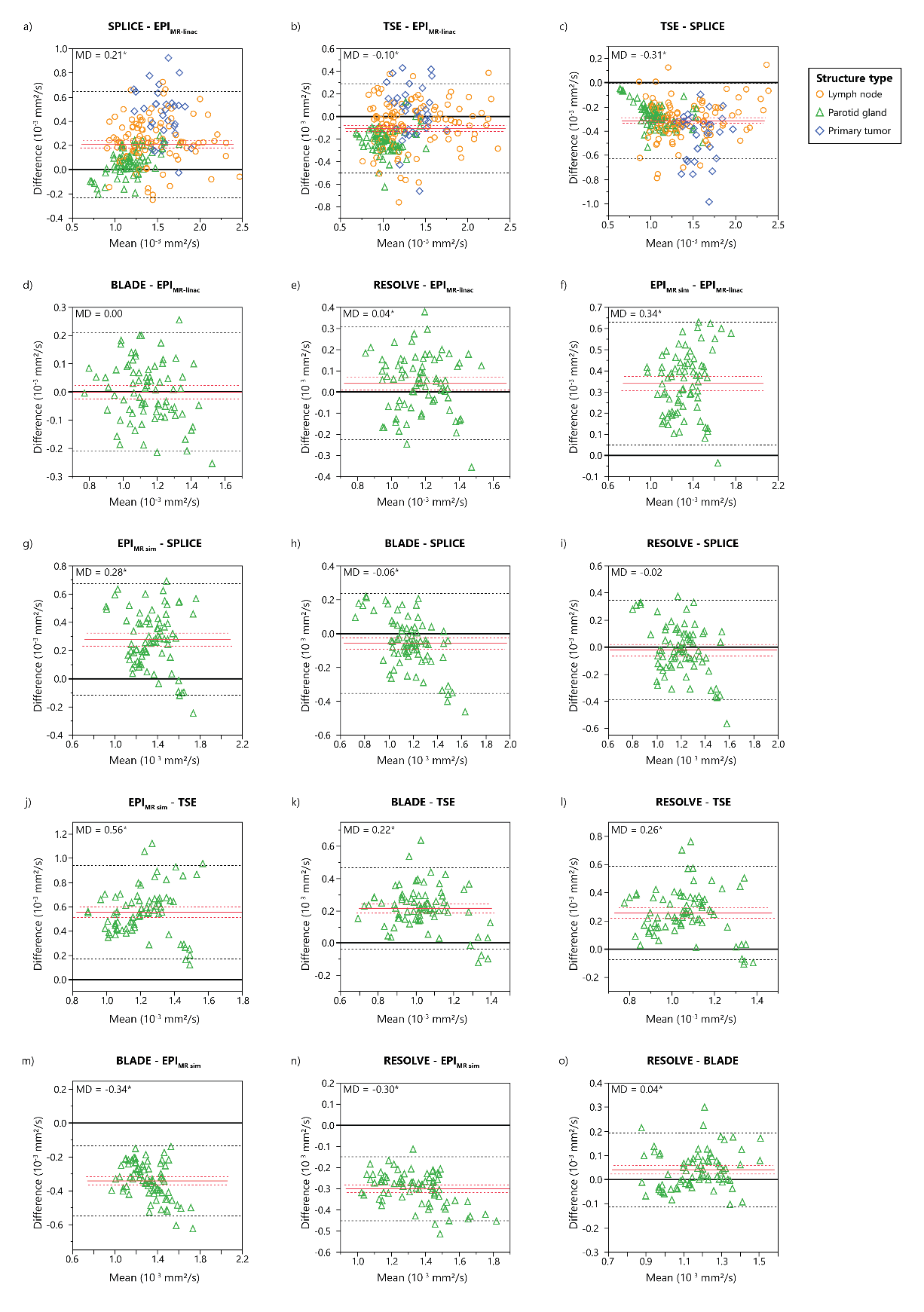

Figure S1: Bland-Altman plots showing differences in measured ADC values between each pair of the three MR-linac DWI sequences (EPI, SPLICE, TSE) and three MR sim DWI sequences (EPI, BLADE, RESOLVE). ADC values represent the mean ADC value within each structure for lymph nodes and primary tumors (patients) and parotid glands (volunteers). Mean differences (MD) are represented by solid red lines with 95% confidence intervals of the MD represented by dotted red lines. 95% limits of agreement are represented by dotted black lines. Statistically significant biases are represented by * (p<0.05).

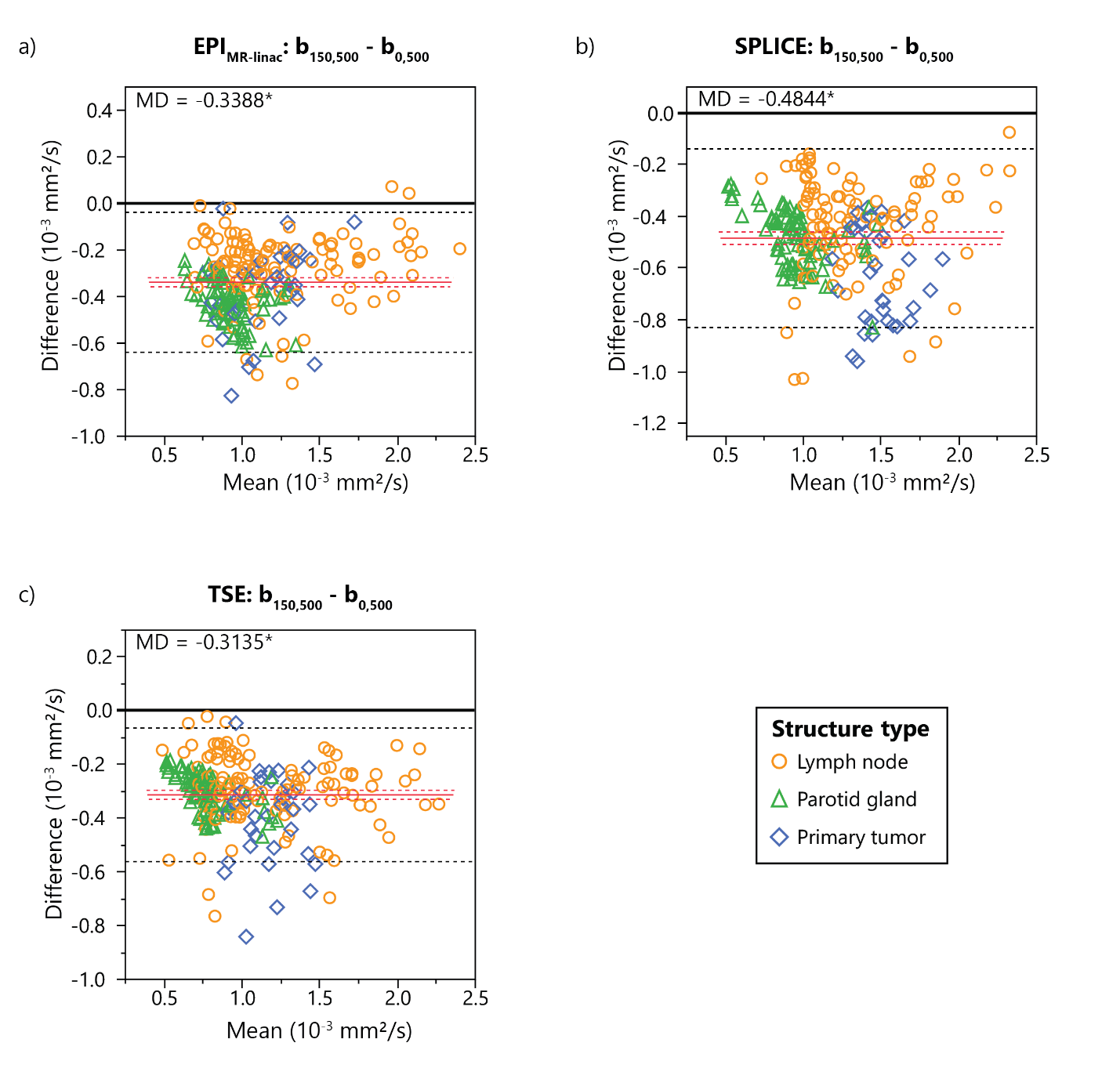

Figure S2: Bland-Altman plots showing differences in measured ADC values between ADC maps reconstructed using b-values of 0 and 500 s/mm^2^ (b_0,500_) and b-values of 150 and 500 s/mm^2^ (b_150,500_) for each of the three MR-linac DWI sequences (EPI, SPLICE, TSE). ADC values represent the mean ADC value within each structure for lymph nodes and primary tumors (patients) and parotid glands (volunteers). Mean differences (MD) are represented by solid red lines with 95% confidence intervals of the MD represented by dotted red lines. 95% limits of agreement are represented by dotted black lines. Statistically significant biases are represented by * (p<0.05).

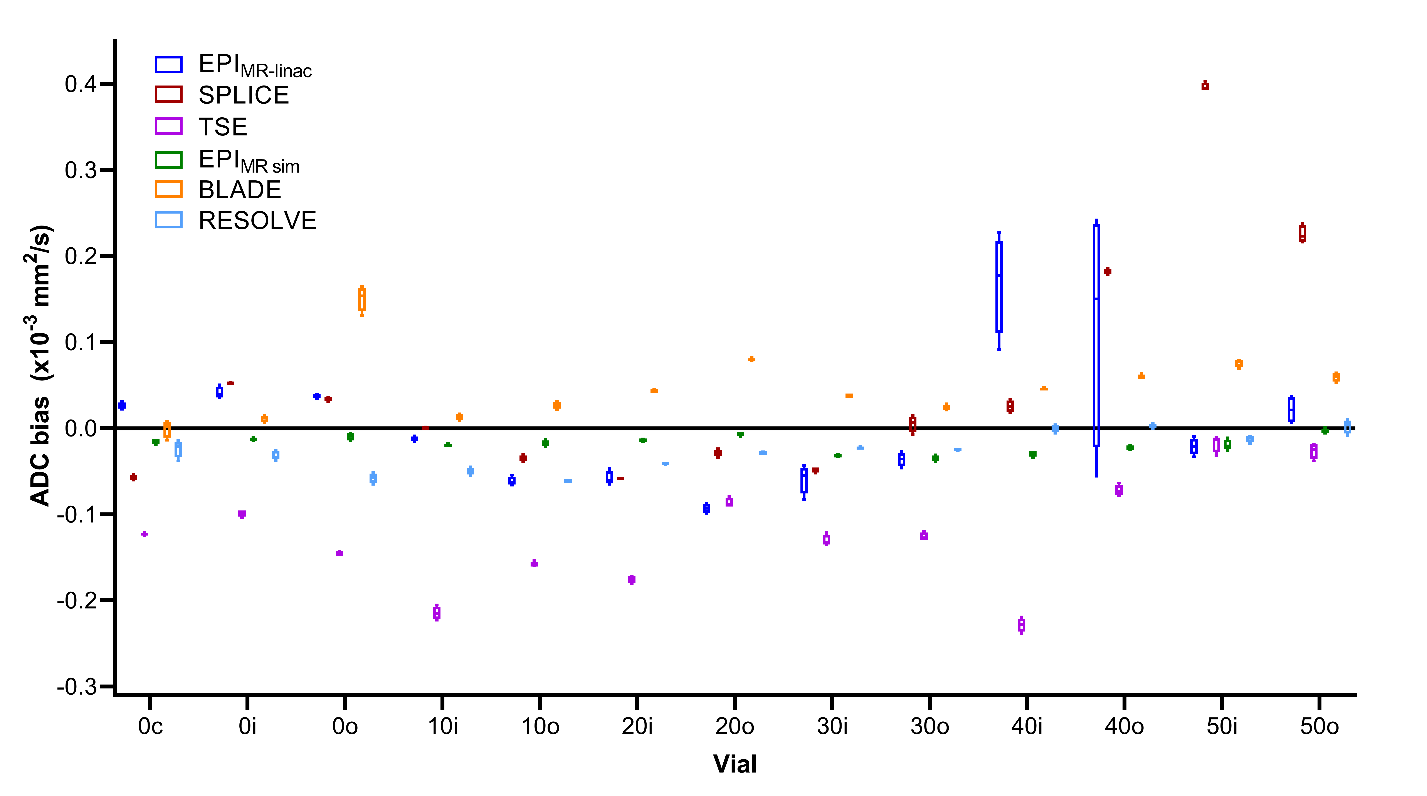
Figure S3: Phantom ADC bias results from four replicate image acquisitions for each individual phantom vial. Each data point within each box-and-whisker plot represents the difference between the mean ADC value in an ROI and the true ADC value for each phantom vial. The numbers in the phantom vial names refer to the concentrations of polymer, which correspond to the manufacturer-provided true ADC value (1.127x10^-3^ mm^2^/s for 0, 0.843x10^-3^ mm^2^/s for 10, 0.607x10^-3^ mm^2^/s for 20, 0.403x10^-3^ mm^2^/s for 30, 0.248x10^-3^ mm^2^/s for 40, and 0.128x10^-3^ mm^2^/s for 50). The letters in the vial names refer to the spatial location within the vial (“c” for “center,” “i” for “inner,” and “o” for “outer”).

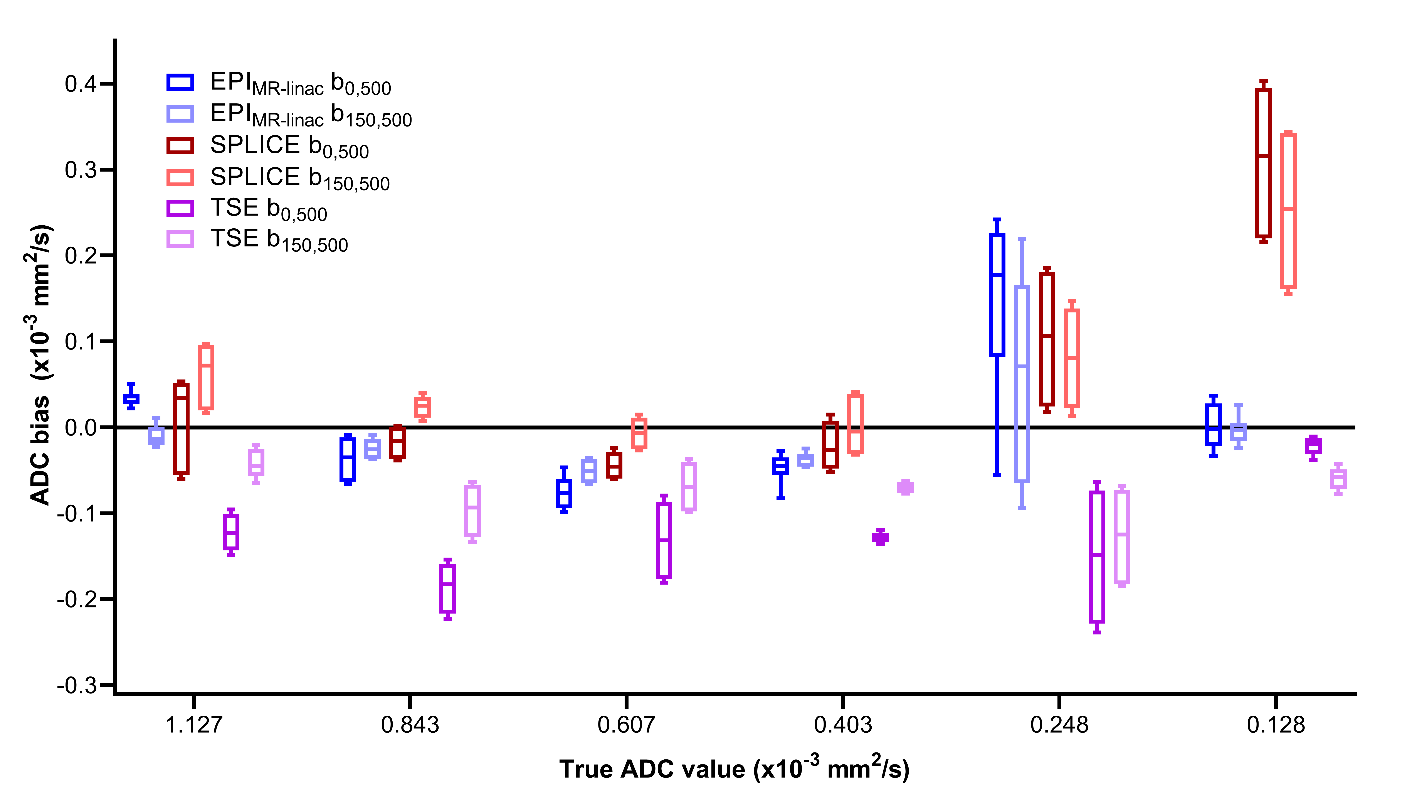

Figure S4: Phantom ADC bias results from four replicate image acquisitions for each MR-linac DWI sequence with ADC maps reconstructed using b-values of 0 and 500 s/mm^2^ (b_0,500_) and b-values of 150 and 500 s/mm^2^ (b_150,500_). Each data point represents the difference between the mean ADC value in an ROI and the true ADC value for each phantom vial. Data points from multiple vials with the same true ADC value are combined.

Table S4: Phantom repeatability and reproducibility metrics for each MR-linac DWI sequence between ADC maps reconstructed using b-values of 0 and 500 s/mm^2^ (b_0,500_) and b-values of 150 and 500 s/mm^2^ (b_150,500_).

|  | EPI_MR-linac_ b_0,500_ | EPI_MR-linac_ b_150,500_ | SPLICE b_0,500_ | SPLICE b_150,500_ | TSE b_0,500_ | TSE b_150,500_ |
| --- | --- | --- | --- | --- | --- | --- |
| %CV (within-ROI) | 2.55 ± 0.71 | 2.15 ± 0.31 | 3.14 ± 0.11 | 2.62 ± 0.06 | 3.62 ± 0.30 | 2.99 ± 0.18 |
| RC (x10^-6^ mm^2^/s) | 10.65 | 4.60 | 8.15 | 5.34 | 4.07 | 7.59 |
| Repeatability wCV (%) | 0.33 | 0.15 | 0.28 | 0.17 | 0.15 | 0.25 |
| RDC (x10^-6^ mm^2^/s) | 30.50 | 6.15 | 25.38 | 16.01 | 2.65 | 0.30 |
| Reproducibility wCV (%) | 0.96 | 0.20 | 0.85 | 0.50 | 0.10 | 0.01 |
